# Assessing the Impact of Mask Mandates on SARS-CoV-2 Transmission: A Case Study of Utah

**DOI:** 10.1101/2023.11.13.23298464

**Authors:** Alicia C. Horn, Holly E. Shoemaker, Lindsay T. Keegan

## Abstract

**Background:** Throughout the COVID-19 pandemic, the effectiveness of face masks mandates has been intensely debated. Many methods have been used to demonstrate mask effectiveness, including one that compares the change in reproduction number following implementing and removing face mask mandates^1^.

**Methods:** Using data from Utah, we calculated the effect of mask mandates (E_Fm_) in each local health district from before and after three key mandates: the Salt Lake and Summit County (SLSC) mask mandates enacted; the Utah statewide mask mandate enacted; and the Utah statewide mandate was lifted.

**Results:** We found that most counties had a reduction in the growth rate of cases following the mandates. There were reductions in E_Fm_ in many counties after the introduction of the SLSC mask mandates and a more widespread reduction in E_Fm_ across the state following the statewide mandate. Lifting the mandates, many counties across the states saw an increase in E_Fm_.

**Conclusion:** Our data show mask mandates were an effective way to reduce transmission both within the jurisdiction they were enacted and in neighboring jurisdictions. We provide evidence to support mask mandates as a way to prevent transmission to be better equipped to respond to future pandemics.

## Introduction

When vaccines and other therapeutics are not available, public health decision makers must rely on non-pharmaceutical interventions (NPI) as community mitigation strategies such as school closures, social distancing, and face mask mandates to reduce disease transmission and severe outcomes^2^. During the COVID-19 pandemic, vaccines against SARS-CoV-2 did not become available until December 2020^3^ consequently, for the first year of the pandemic, all personal and community mitigation efforts came in the form of NPIs.

While numerous NPIs were considered, mask mandates became the primary approach to slowing the spread of COVID-19^4^. Face masks have been utilized as a method to slow transmission before the start of the COVID-19 pandemic, especially in other countries during flu season^5^. However, masks have been the catalysts of intense debates fueled by politics, with different states choosing to implement and enforce mandates often based on their political leanings^6,7^.

The deployment of mask mandates in Utah is representative of the fragmented nature of the way that interventions were employed throughout the state. The first mandates applied in the state occurred in both Salt Lake and Summit counties (SLSC), which received approval on June 27th, 2020. The next mandate was put in place in Grand County on July 7th, 2020. As cases began to spike in the fall of 2020, there were scattered attempts from different health departments to deal with the rising cases, creating a mosaic of interventions throughout the state ranging from recommendations to actual mandates. Throughout this period, there were special mandates for schools and public transportation^8,9^. A statewide mandate was issued on November 9th, 2020 for all counties until April 10th, 2021. After the statewide mandate ended, Salt Lake City opted to keep their mandates longer. We show a labeled map of the Local Health Districts (LHDs) in the supplement (**Figure SI 1**).

Many studies measure the efficacy of mask mandates on the spread of airborne diseases which show masks to be effective at protecting individuals from infection^1,10–20^. One method, described by Britton et al.^1^ estimates the effect of mask mandates at the population level by comparing the change in the growth rate of cases before and after a mandate is implemented or lifted.

One benefit of the fragmented system of mandates in Utah is that it has created a natural experiment to compare the efficacy of these different interventions within the same state by comparing the data between health districts. Here, we show how the state, county, and local mandates differed in their effectiveness in reducing the number of COVID-19 cases within the jurisdiction where the mandate was implemented as well as across the state of Utah.

## Methods

The data for this project is from the Utah Department of Health and Human Services COVID-19 surveillance dashboard^21^.

We employ the method outlined in Britton et al.^1^ to calculate the *Face Mask Effect* (E_Fm_). E_Fm_ is calculated by comparing the effective reproduction number (R_e_) pre- and post-intervention, and is given by the following equation:

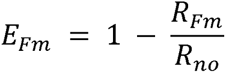

Where R_Fm_ and R_no_ are calculated by:

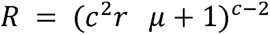

Additionally, *r* is calculated before (*r_no_*) or after (*r_Fm_*) a mask mandate is implemented, and is given by:

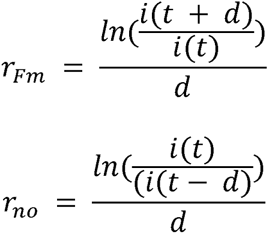

In this study, we consider 28 days before and after the mask mandate to account for the variabilities in Utah’s reported cases with respect to day of the week due to weekend lab closures.

We calculated the E_Fm_ at the at the LHD level across the state following three dates: June 28th, 2020 the date of the SLSC mask mandates^23,24^ November 9th, 2020 the date of the Utah State-wide mask mandate^25,26^ and April 10th, 2021 the date when all mandates were lifted except in Salt Lake City^27^. All analyses were conducted in R^28^, all code is available on github^29^.

## Results

Mask mandates were implemented in Utah at times when cases were trending upwards and there was concern for healthcare infrastructure (**Figure 1**). Following the SLSC mask mandate on June 28, 2020, Salt Lake County Health District (HD) saw a 22.4% reduction in R_e_ in the 28 days following the mandate compared to the 28 days preceding the mandate (**Table 2**, **Figure 2**). Conversely, Summit County HD saw a 6.7% increase in R_e_ following the mandate (**Table 2**, **Figure 2**). After the statewide mandate was implemented on November 9, 2020, all of the LHDs across the state except San Juan HD saw reductions in R_e_ following the mandate (**Table 2**, **Figure 2**). After the statewide mandate was lifted on April 10, 2021 six LHDs exhibited a rise in cases, with some more gradual than others (**Table 2**, **Figure 2**). Southeast Utah HD had the largest increase in R_e_ (26.2%) and Salt Lake County HD had the smallest increase of 1.9% following lifting the mandate.

**Figure 1:**
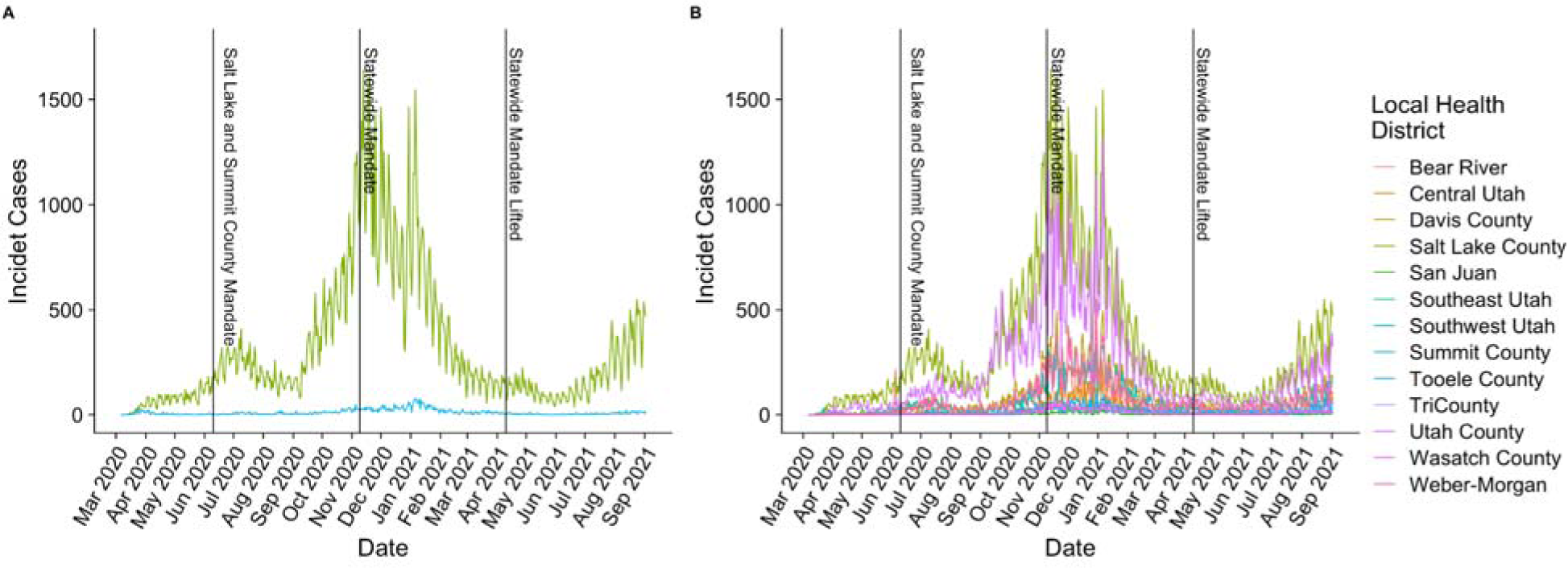
Plot of incident cases of SARS-CoV-2 by county health district over the course of the COVID-19 pandemic with vertical lines indicating the implementation and lifting of the different mask mandates as labeled. **A)** Incident cases of SARS-CoV-2 in SLSC and **B)** Incident cases of SARS-CoV-2 in all Health Districts in Utah.

**Figure 2:**
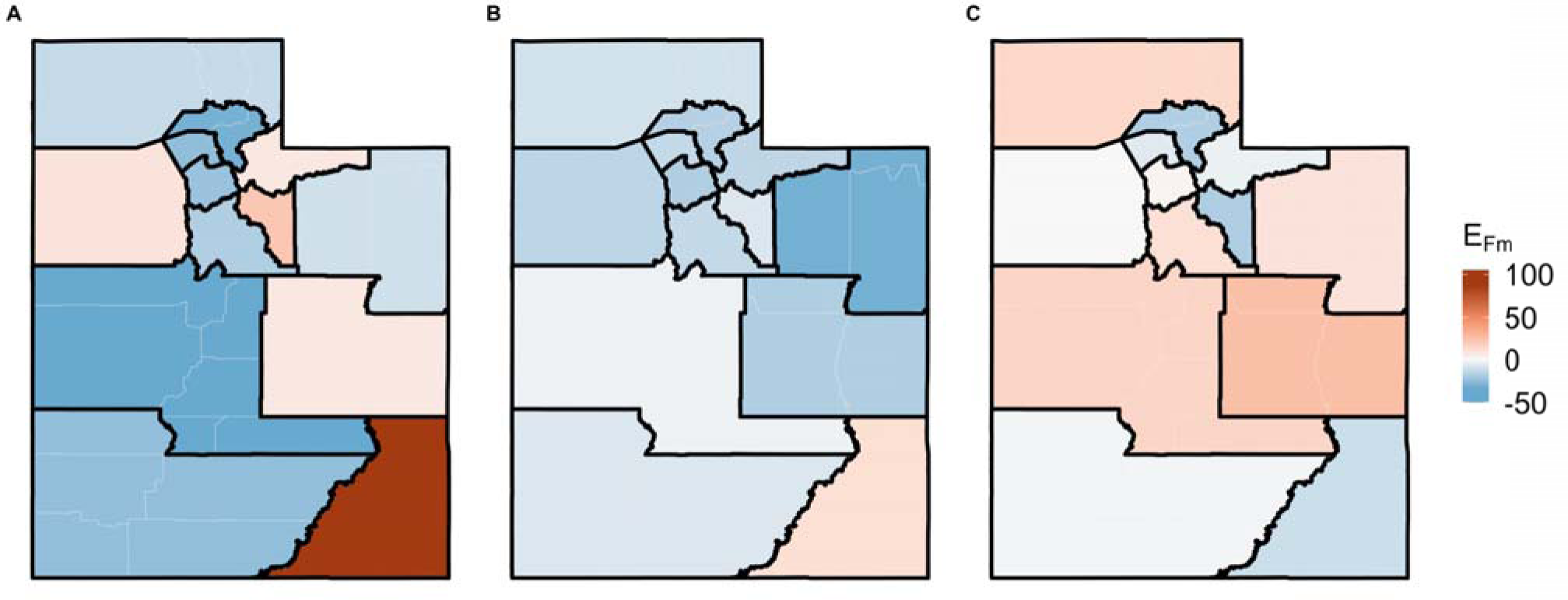
Plot of the Effect of Mask Mandates (E_Fm_) following the implementation or removal of mask mandates. Where a negative E_Fm_ corresponds to a reduction in transmission while a positive E_Fm_ corresponds to an increase in transmission following the mandate change. A) Shows the effect following the implementation of the SLSC mask Mandates on June 27, 2020, B) shows the effect following the implementation of the statewide mandate on November 9, 2020, and C) shows effect of lifting the statewide mandate on April 10th, 2021.

**Table 1:** Tables of variables, their meaning, and if applicable the value we used for this study.

| Variable | Meaning | Value |
| --- | --- | --- |
| $E_{Fm}$ | Effectiveness of mask mandates | Estimated |
| $R_{Fm}$ | Effective reproduction number following the implementation of a mask mandate | Estimated |
| $R_{no}$ | the effective reproduction number prior to the implementation of a mask mandate | Estimated |
| $\mu$ | Mean generation time | 6.5 days<sup>22</sup> |
| $\sigma$ | Standard deviation of the generation time distribution | 4 days<sup>22</sup> |
| $c = \frac{\sigma}{\mu}$ | Coefficient of variation of the generation time distribution | 1.625<sup>22</sup> |
| $r$ | The growth rate of cases | Estimated |
| $d$ | Number of days before or after a mandate that we consider. | 28 days |
| $r_{no}$ | the growth rate of cases $d$ days before a mask mandate is implemented | Estimated |
| $r_{Fm}$ | the growth rate of cases $d$ days after a mask mandate is implemented | Estimated |
| $i(t)$ | The incidence at the time of the mandate (t) | Estimated |

**Table 2:** Table of the effectiveness of mandates implementing or lifting masking requirements by LHD in Utah for the SLSC mask mandate on 6/28/2020, the statewide mask mandate on 11/9/2020, and the lifting of the statewide mask mandate on 4/10/2020.

| Health District | $E_{Fm}$ for SLSC mask mandate | $E_{Fm}$ for statewide Mask Mandate | $E_{Fm}$ for the lifting of the statewide Mask Mandate* |
| --- | --- | --- | --- |
| Salt Lake County Health District* | -22.4% | -17.2% | 1.9% |
| Summit County Health District | 6.68% | -14.2% | -2.9% |
| Wasatch County Health District | 22.0% | -7.0% | -17.8% |
| Weber-Morgan Health District | -30.2% | -16.2% | -17.1% |
| TriCounty Health District | -9.9% | -30.8% | 9.5% |
| Southwest Utah Health District | -24.3% | -7.2% | -1.4% |
| Southeast Utah Health District | 6.9% | -17.4% | 26.2% |
| San Juan Health District | 104.5% | 11.2% | -11.0% |
| Central Utah Health District | -42.4% | -2.4% | 15.6% |
| Tooele County Health District | 9.1% | -14.4% | -0.04% |
| Davis County Health District | -24.6% | -12.5% | -5.7% |
| Utah County Health District | -17.1% | -12.7% | 11.0% |
| Bear River Health District | -11.9% | -9.3% | 14.4% |

\*Salt Lake City still had a mandate but the county as a whole did not

In addition to the mandates impacting the jurisdictions for which they were levied, the SLSC mandates had an impact on other LHDs across the state (**Table 2**, **Figure 2**). The largest effect was in the Central Utah HD which had a 42.4% reduction in R_e_, but LHDs along the interstate highway I-15 corridor all saw a reduction in R_e_. However, not all LHDs saw a reduction following the SLSC mask mandates, 5 counties saw an increase in R_e_ and the LHD with the highest increase was San Juan HD which had a 104.5% increase in R_e_ following the mandate (**Table 2**).

## Discussion

Overall we found that the majority of counties experienced a reduction in transmission after mask mandates were implemented. The most comprehensive reductions were shown after the statewide mandate was implemented with nearly all health districts exhibiting some decrease (except San Juan HD). After the statewide mandate was lifted, most LHDs exhibited an increase in transmission. While the statewide mandate was lifted in April 2020, Salt Lake City maintained a city-wide mask mandate until 2022, which may partially explain why Salt Lake County HD only saw a modest increase in transmission.

Although it is not surprising that mask mandates resulted in reductions in transmission within the jurisdiction that they were issued, we found a surprising trend that many additional LHDs exhibited a reduction in transmission following a mask mandate in a different jurisdiction. In particular, we found that LHDs that run along the interstate highway I-15 corridor all exhibited a reduction in transmission following the SLSC mask mandate. This suggests that mandates in large population hubs like Salt Lake County can have far reaching impacts. This could indicate that people may be changing individual behavioral patterns in response to other jurisdictions mandates and rising case counts.

Another interesting finding was that the largest reductions in transmission occurred after the implementation of the June 2020 mandates with most LHDs exhibiting a larger decrease following these mandates than after the statewide mandate. The statewide mandate reduced transmission across all LHDs (except one) including counties that did not exhibit a decrease following the SLSC mandates. One potential explanation for this reduced impact could be that counties throughout the state had already begun to apply their own mask restrictions in the weeks leading up to the statewide mandate or that individuals had already started to change their own behavior in the absence of mandates as cases of COVID-19 were increasing at that time.

Exploring the patterns of counties that did not see an impact of mask mandates can also be illuminating. There was an increase in transmission in Summit County following the SLSC mandate, which could be attributed to the fact that their case levels were very low when they instituted the mandate, averaging 39.1 cases per week prior to the mandate and 112.7 cases per week after. Additionally, San Juan HD saw a 104.5% increase in R_e_ following the SLSC mask mandates, and a 11.2% increase in R_e_ following the statewide mandate. One potential explanation for this is that 21% of San Juan HD’s population is self identified as Navajo^30^ and the Navajo nation was experiencing a major outbreak of SARS-CoV-2 during this time^30,31^.

Our findings are particularly interesting when taken in the broader context of mandates. Seegert et al.^32^ explored the economic impacts of mask mandates and found that statewide mandates stimulate economic activity while county-wide mandates depress activity, suggesting that this could be a result of risk perception^32^. Taken with our findings that mask mandates generally reduced transmission within the jurisdiction where levied as well as in connected jurisdictions, it suggests that in future disease outbreak scenarios, swift, statewide action may be equally effective at reducing transmission and more effective at reducing economic harms than a piecemeal, local approach.

A number of studies have attempted to estimate the effectiveness of face mask mandates^1,10–20^. One randomized control trial concluded that wearing masks reduced the risk of infection for the wearer by 18%; while another found the symptomatic incidence was reduced 9.5%^10,11^. Observational studies that directly estimate the effect of mask mandates on R_e_,estimated a 29% reduction in Kansas^1,15^, a 21–43% reduction in Jena, Germany^1,17^, and a 15.1% reduction when averaged over 190 countries and adjusted for confounders^16^. Further, a simulation-based study found masking equated to an average 25-35% reduction in infectious contacts^13^. Overall, our study shows a similar median effectiveness of 12.7% reduction following the implementation of the statewide mandate across all LHDs (a 13.5% reduction when excluding San Juan HD) and a similar 11.9% reduction across all LHDs following the implementation of the SLSC mandate (a 7.9% reduction when just considering the LHDs with mandates).

Our study attempts to evaluate the effectiveness of mask mandates as they are implemented. However, it has several limitations. We use government mask mandates as a proxy for mask wearing. Unfortunately, the relationship between mandates and mask use is relatively weak, with one study showing no statistically significant relationships and other studies showing between a 13% – 23% relationship between mandates and mask use in the United States^18,19,33^. Consequently, just because mandates were in place, doesn’t mean that there was full compliance. Many mandates were designed to be lightly enforced, the tense political climate surrounding mask use reduced willingness to enforce mask mandates, potentially decreasing the efficacy of those mandates^34^. Further, some people may have worn a mask regardless of mandate status. Our study does not attempt to link actual mask usage to the implementation of masking mandates.

Here, we examine mask mandates and their impact at the population-level. While mask mandates dictate that individuals must wear a mask, many people used ineffective masks^35^. Our study does not attempt to provide evidence on the individual effectiveness of using a mask.

Another potential limitation of our study is the timing of vaccine availability. Vaccines first started becoming available to healthcare workers in Utah in December 2020,becoming widely available to the general public by March 2021^3,36,37^. Given the timing of the statewide mask mandate on November 9, 2020, we do not expect that vaccination impacted estimates of the face mask effectiveness. However, with widespread vaccination access beginning in March 2021, it is possible that vaccination may have impacted estimates of the effectiveness of removing the statewide mandate in April 2021. However, we believe that this impact was limited as only 32% of people in Utah were fully vaccinated by May 8th, 2021 which is when our study’s time frame ends^38^.

## Conclusions

Our study shows that the implementation of mask regulations in Utah played a role in reducing SARS-CoV-2 transmission throughout the state. We found that implementing and lifting mask mandates influenced both the jurisdictions under which the mandate was levied as well as connected jurisdictions. The COVID-19 pandemic is unlikely to be the last time we are faced with the transmission of an airborne, viral pathogen; consequently, understanding the most effective way to reduce transmission is essential to inform future pandemic response.

## Data Availability

The data for this project are publicly available from the Utah Department of Health and Human Services COVID-19 surveillance dashboard.

https://coronavirus-dashboard.utah.gov/overview.html

## Acknowledgements

ACH and LTK acknowledge funding provided by the Centers for Disease Control and Prevention (75D30121F00003). HES and LTK acknowledge funding provided by the Centers for Disease Control and Prevention (1U01CK000585-01). The contents are solely the responsibility of the authors and do not necessarily represent the official position or policy of the Centers for Disease Control and Prevention. ACH acknowledges funding provided by the Undergraduate Research Opportunities Program at the University of Utah awarded to Alicia C. Horn.

---

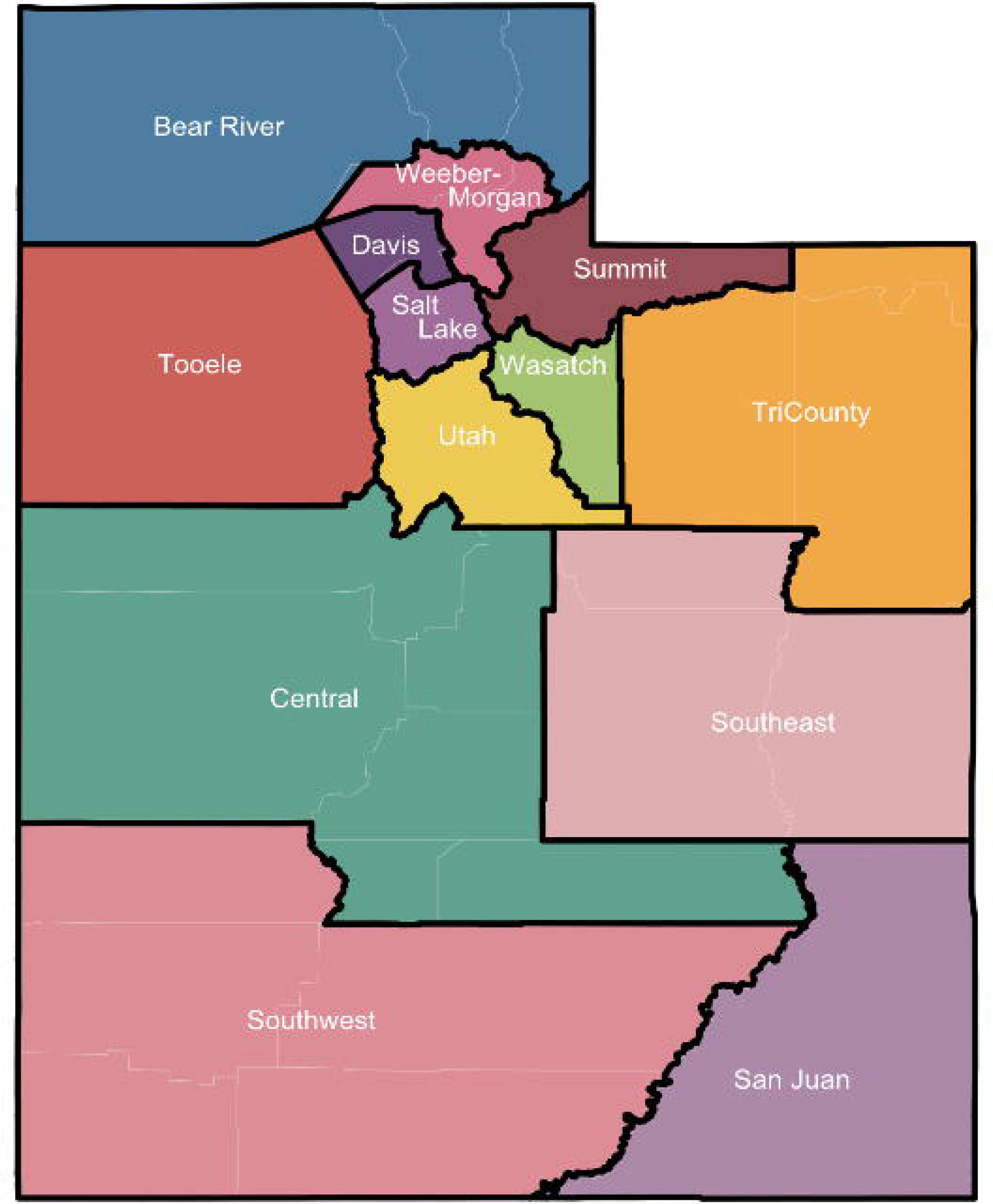

## References

1. Britton T. Quantifying the preventive effect of wearing face masks. Proc Math Phys Eng Sci. Jul 2021;477(2251):20210151. doi:10.1098/rspa.2021.0151

2. Lai S, Ruktanonchai NW, Zhou L, et al. Effect of non-pharmaceutical interventions to contain COVID-19 in China. Nature. 2020/09/01 2020;585(7825):410–413. doi:10.1038/s41586-020-2293-x

3. Stevens T. Gov. Spencer Cox says all Utahns 16 and older can start making COVID-19 vaccine appointments next week. The Salt Lake Tribune. https://www.sltrib.com/news/politics/2021/03/18/watch-live-gov-spencer/

4. Krishnamachari B, Morris A, Zastrow D, Dsida A, Harper B, Santella AJ. The role of mask mandates, stay at home orders and school closure in curbing the COVID-19 pandemic prior to vaccination. American journal of infection control. 2021/08/01/ 2021;49(8):1036–1042. 10.1016/j.ajic.2021.02.002

5. Bish A, Michie S. Demographic and attitudinal determinants of protective behaviours during a pandemic: A review. British Journal of Health Psychology. 2010;15(4):797–824. 10.1348/135910710X485826

6. Rains SA, Colombo PM, Quick BL, Kriss LA. State mask mandates and psychological reactance theory: The role of political partisanship and COVID-19 risk in mask adoption and resistance. Social science & medicine. 2022/12/01/ 2022;314:115479. 10.1016/j.socscimed.2022.115479

7. Adolph C, Amano K, Bang-Jensen B, et al. Governor Partisanship Explains the Adoption of Statewide Mask Mandates in Response to COVID-19. State Politics & Policy Quarterly. 2022;22(1):24–49. doi:10.1017/spq.2021.22

8. Breslow J. CDC Issues Sweeping New Mask Mandate For U.S. Travelers, Extends Eviction Moratorium. npr. https://www.npr.org/sections/coronavirus-live-updates/2021/01/30/962390180/cdc-issues-sweeping-new-mask-mandate-for-u-s-travelers-extends-eviction-moratori

9. Lanier WA, Babitz KD, Collingwood A, et al. COVID-19 Testing to Sustain In-Person Instruction and Extracurricular Activities in High Schools - Utah, November 2020-March 2021. MMWR Morbidity and mortality weekly report. May 28 2021;70(21):785–791. doi:10.15585/mmwr.mm7021e2

10. Effectiveness of Adding a Mask Recommendation to Other Public Health Measures to Prevent SARS-CoV-2 Infection in Danish Mask Wearers. Annals of internal medicine. 2021;174(3):335–343. doi:10.7326/m20-6817%m33205991

11. Abaluck J, Kwong LH, Styczynski A, et al. Impact of community masking on COVID-19: A cluster-randomized trial in Bangladesh. Science. 2022;375(6577):eabi9069. doi:doi:10.1126/science.abi9069

12. Cash-Goldwasser S, Reingold AL, Luby SP, Jackson LA, Frieden TR. Masks During Pandemics Caused by Respiratory Pathogens—Evidence and Implications for Action. JAMA Network Open. 2023;6(10):e2339443–e2339443. doi:10.1001/jamanetworkopen.2023.39443

13. Koslow W, Kühn MJ, Binder S, et al. Appropriate relaxation of non-pharmaceutical interventions minimizes the risk of a resurgence in SARS-CoV-2 infections in spite of the Delta variant. PLOS Computational Biology. 2022;18(5):e1010054. doi:10.1371/journal.pcbi.1010054

14. Flaxman S, Mishra S, Gandy A, et al. Estimating the effects of non-pharmaceutical interventions on COVID-19 in Europe. Nature. 2020/08/01 2020;584(7820):257–261. doi:10.1038/s41586-020-2405-7

15. Van Dyke ME, Rogers TM, Pevzner E, et al. Trends in County-Level COVID-19 Incidence in Counties With and Without a Mask Mandate - Kansas, June 1-August 23, 2020. MMWR Morbidity and mortality weekly report. Nov 27 2020;69(47):1777–1781. doi:10.15585/mmwr.mm6947e2

16. Bo Y, Guo C, Lin C, et al. Effectiveness of non-pharmaceutical interventions on COVID-19 transmission in 190 countries from 23 January to 13 April 2020. International Journal of Infectious Diseases. 2021/01/01/ 2021;102:247–253. 10.1016/j.ijid.2020.10.066

17. Mitze T, Kosfeld R, Rode J, Wälde K. Face masks considerably reduce COVID-19 cases in Germany. Proceedings of the National Academy of Sciences of the United States of America. Dec 22 2020;117(51):32293–32301. doi:10.1073/pnas.2015954117

18. Leech G, Rogers-Smith C, Monrad JT, et al. Mask wearing in community settings reduces SARS-CoV-2 transmission. Proceedings of the National Academy of Sciences of the United States of America. Jun 7 2022;119(23):e2119266119. doi:10.1073/pnas.2119266119

19. Rader B, White LF, Burns MR, et al. Mask-wearing and control of SARS-CoV-2 transmission in the USA: a cross-sectional study. Lancet Digit Health. Mar 2021;3(3):e148–e157. doi:10.1016/s2589-7500(20)30293-4

20. Maloney MJ, Rhodes NJ, Yarnold PR. Mask mandates can limit COVID spread: Quantitative assessment of month-over-month effectiveness of governmental policies in reducing the number of new COVID-19 cases in 37 US States and the District of Columbia. medRxiv. 2020:2020.10.06.20208033. doi:10.1101/2020.10.06.20208033

21. Overview of COVID-19 Surveillanc. https://coronavirus-dashboard.utah.gov/overview.html

22. Chen D, Lau Y-C, Xu X-K, et al. Inferring time-varying generation time, serial interval, and incubation period distributions for COVID-19. Nature Communications. 2022/12/13 2022;13(1):7727. doi:10.1038/s41467-022-35496-8

23. Johnson E. Salt Lake and Summit Counties mask mandate to take effect. ABC4 News. https://www.abc4.com/coronavirus/salt-lake-and-summit-counties-mask-mandate-take-effect/

24. Public Health Order. https://slco.org/globalassets/1-site-files/health/programs/covid/pho/pho11.pdf

25. Salcedo A. Utah govenor declares emergencey issues mask mandate ‘We can not afford to debate this issue’. The Washington Post. https://www.washingtonpost.com/nation/2020/11/09/utah-emergency-masks-mandate-covid/

26. Governor Declares New State of Emergency to Address Hospital Overcrowding, Case Surge https://coronavirus-download.utah.gov/Governor/State-of-emergency-press-release-11.8.2020.pdf

27. UTAH’S STATEWIDE MASK MANDATE ENDS TODAY. HERE’S WHAT YOU NEED TO KNOW. https://governor.utah.gov/2021/04/10/utahs-statewide-mask-mandate-ends-today-heres-what-you-need-to-know/

28. R: A language and environment for statistical computing. https://www.R-project.org

29. Utah ID Dyanmics. https://github.com/UT-IDDynamics

30. Denetclaw WF, Otto ZK, Christie S, et al. Diné Navajo Resilience to the COVID-19 pandemic. PloS one. 2022;17(8):e0272089. doi:10.1371/journal.pone.0272089

31. Navajo Epidemiology Center Helping with Making Data-driven Decisions for the COVID-19 response. https://nec.navajo-nsn.gov/Portals/0/Home%20Webpage/NEC.COVID-19.TwoPager.Oct29.20.v1e.PRINT.pdf?ver=uD5B7e_rWbnuDywbmWonGQ%3d%3d&timestamp=1629923663010

32. Seegert N, Gaulin M, Yang M-J, Navarro-Sanchez F. Information revelation of decentralized crisis management: Evidence from natural experiments on mask mandates. Available at SSRN 3736407. 2020;

33. Dhaval Adjodah KD, Matteo Chinazzi, Samuel P. Fraiberger, Alex Pentland, Samantha Bates, Kyle Staller, Alessandro Vespignani, Deepak L. Bhatt. Association between COVID-19 Outcomes and Mask Mandates, Adherence, and Attitudes. 2020;

34. Lyons J, Fowler L. Is It Still a Mandate If We Don’t Enforce It? The Politics of COVID-related Mask Mandates in Conservative States. State and Local Government Review. 2021;53(2):106–121. doi:10.1177/0160323x211035677

35. Chu DK, Akl EA, Duda S, Solo K, Yaacoub S, Schünemann HJ. Physical distancing, face masks, and eye protection to prevent person-to-person transmission of SARS-CoV-2 and COVID-19: a systematic review and meta-analysis. Lancet. Jun 27 2020;395(10242):1973–1987. doi:10.1016/s0140-6736(20)31142-9

36. Mathieu E, Ritchie H, Ortiz-Ospina E, et al. A global database of COVID-19 vaccinations. Nature Human Behaviour. 2021/07/01 2021;5(7):947–953. doi:10.1038/s41562-021-01122-8

37. Walker C. Gov Cox: All Utah residents eligible for vaccine next Wednesday. KSLNewsRadio. https://kslnewsradio.com/1945307/all-utahns-eligible-for-vaccine-next-wednesday/

38. Leonard W. The next group eligible for COVID-19 vaccination in Utah could help further curb transmission. DeseretNews. https://www.deseret.com/utah/2021/5/8/22426318/the-next-group-eligible-for-covid-19-vaccination-in-utah-could-help-further-curb-transmission

