## Supplemental Materials for "Assessing the Impact of Mask Mandates on SARS-CoV-2 Transmission: A Case Study of Utah"

**Supplemental Information**

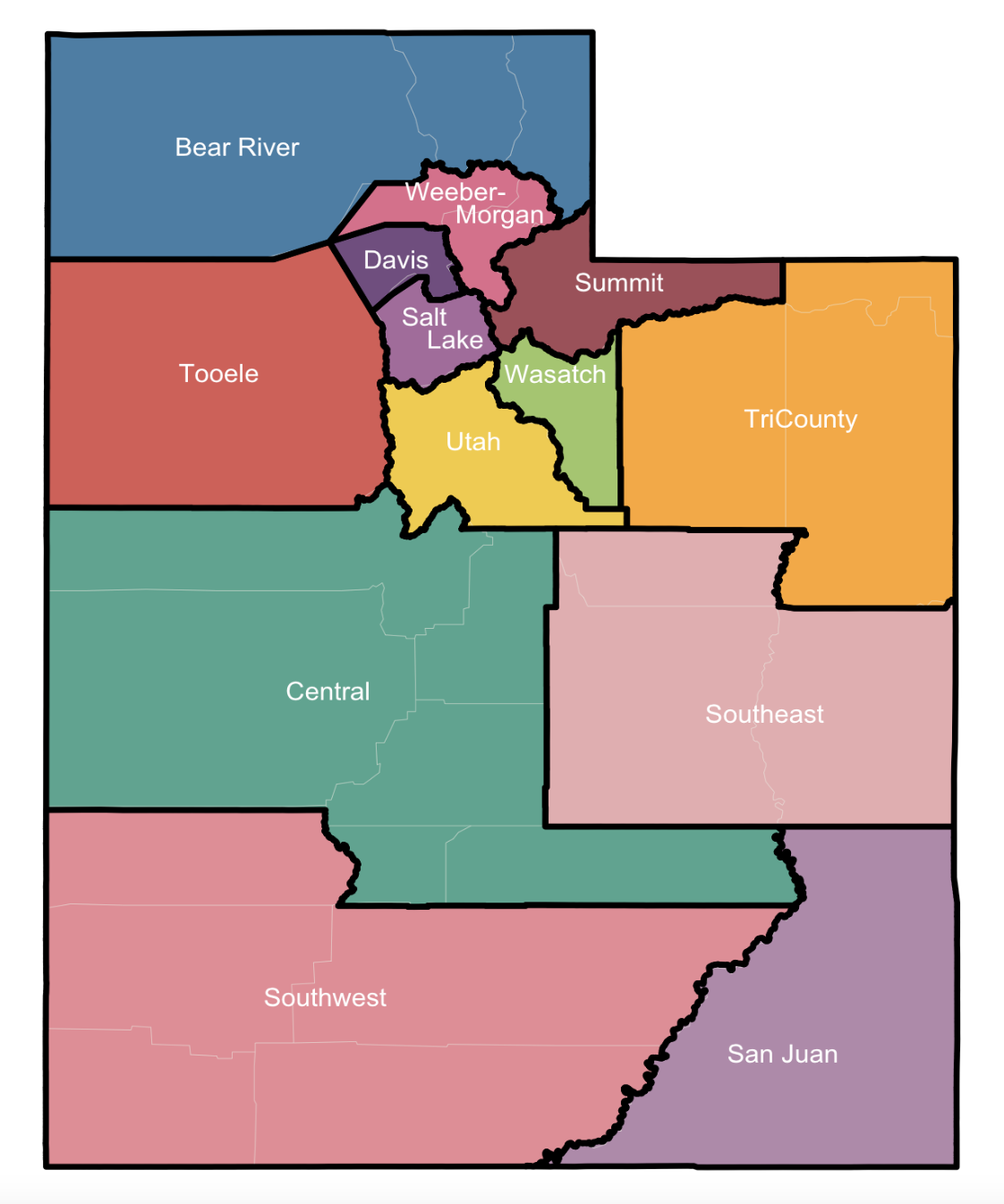

**Figure SI 1: Labeled map of the LHDs in Utah**
